# Initial Antidepressant Choice by Non-Psychiatrists: Learning from Large-scale Electronic Health Records

**DOI:** 10.1101/2021.08.02.21260076

**Authors:** Yi-han Sheu, Colin Magdamo, Matthew Miller, Jordan W. Smoller, Deborah Blacker

## Abstract

**Introduction:** Pharmacological treatment of depression mostly occurs in non-psychiatric settings, but factors that determine the initial choice of antidepressant treatment in these settings are not well-understood. This study models how non-psychiatrists choose among four antidepressant classes at first prescription (selective serotonin reuptake inhibitors [SSRI], bupropion, mirtazapine, or serotonin-norepinephrine reuptake inhibitors [SNRI]), by analyzing electronic health record (EHR) data.

**Methods:** EHR data were from the Mass General Brigham Healthcare System (Boston, Massachusetts, USA) for the period from 1990 to 2018. From a literature search and expert consultation, we selected 64 variables that may be associated with antidepressant choice. Patients who participated in the study were aged 18 to 65 at the time of first antidepressant prescription with a co-occurring International Classification of Diseases (ICD) code for a depressive disorder. Multinomial logistic regression with main effect terms for all 64 variables was used to model the choice of antidepressant. Using SSRI as the reference class, odds ratios, 95% confidence intervals (CI), and likelihood ratio-based p-values for each variable were reported. We used a false discovery rate (FDR) with the Benjamini–Hochberg procedure to correct for multiple comparisons.

**Findings:** A total of 47,107 patients were included after application of inclusion/exclusion criteria. We observed significant associations for 36 of 64 variables after multiple comparison corrections. Many of these associations suggested that antidepressants’ known pharmacological properties/actions guided choice. For example, there was a decreased likelihood of bupropion prescription among patients with epilepsy (adjusted OR 0.41, 95% CI: 0.33–0.51, p < 0.001), an increased likelihood of mirtazapine prescription among patients with insomnia (adjusted OR 1.58, 95% CI: 1.39–1.80, p < 0.001), and an increased likelihood of SNRI prescription among patients with pain (adjusted OR 1.22, 95% CI: 1.11–1.34, p = 0.001).

**Interpretation:** Non-psychiatrists’ selection of antidepressant class appears to be guided by clinically relevant pharmacological properties, indications, and contraindications, suggesting that broadly speaking they choose antidepressants based on meaningful differences among medication classes.

## INTRODUCTION

Major depressive disorder (MDD) is one of the most common psychiatric disorders, with a lifetime prevalence in the U.S. of approximately 20% (1). Pharmacological treatment of depression mostly occurs in non-psychiatric settings (2), although little is known about antidepressant treatment selection practices in primary care, and whether factors affecting treatment decisions in these settings are consistent with expert recommendations (3–6). In this analysis, we take advantage of the comprehensive longitudinal data captured in EHRs to examine the factors associated with antidepressant selection by non-psychiatric clinicians. In addition to using coded (structured) data in EHR, we also applied natural language processing (NLP) to extract mental health symptom information that is documented in free text (unstructured) data from clinical notes. We discuss the extent to which these factors align with those recommended in the APA Practice Guideline and the psychiatric literature (3–6).

Factors considered included patient demographics; comorbidities; depression-related mental symptoms; drug side effects; and drug-drug interactions.

## METHODS

### Institutional Review Board (IRB) approval

This study was approved by the IRB of Mass General Brigham (MGB) Healthcare.

### Data source

Data were extracted from the Research Patient Data Repository (RPDR) (7) of the MGB Healthcare System in Boston, Massachusetts. The RPDR includes data on more than 7 million patients and over 3 billion records from 7 hospitals, including 2 major academic hospitals. Clinical data recorded in the RPDR include encounter (patient visit) meta-data (e.g., time, location, provider, etc.), demographics, ICD 9 and 10 Clinical Modification (ICD-9-CM and ICD-10-CM) diagnoses, laboratory tests, medications, procedures, and patient reported outcome measures.

### Study population

EHR data were extracted for the period January 1990 to August 2018. Patients included were between age 18 and 65 at the time of their first antidepressant prescription (comprising any one of the following: citalopram, escitalopram, fluoxetine, fluvoxamine, paroxetine, sertraline, venlafaxine, desvenlafaxine, duloxetine, mirtazapine, bupropion, vilazodone, and vortioxetine) with a co-occurring diagnostic ICD code for a depressive disorder (defined as ICD-9-CM: 296.20–6, 296.30–6, and 311; ICD-10-CM: F32.1–9, F32.81, F32.89, F32.8, F33.0–3, and F33.8–9). We defined the first visit with co-occurring antidepressant prescription and depression ICD code as the “index visit.” We excluded patients based on the following criteria: (1) first prescription was given prior to 1997, when the latest antidepressant category (mirtazapine) became available; (2) first prescription was made by a psychiatrist, as our goal was to study non-psychiatrist prescribing practices; (3) absence of clinical notes or details of visits within the 90-day window prior to the date of first prescription (to ensure that data were available to address whether the index date really reflected the initial prescription [see below]); (4) presence of ICD codes for bipolar disorder, schizophrenia, or schizoaffective disorder on or prior to the date of first prescription; (5) first documented prescription included two or more antidepressants, as these were unlikely to be first prescriptions; and (6) prescription of an antidepressant other than the four classes of interest.

Because the data are limited to what is recorded in the MGB EHR, some patients may have already been on an antidepressant prior to the index visit (“prevalent users”) due to previous outside prescriptions. For such patients, the pattern of association between the predictors measured at or around the index visit may be different than those of true initiators (“new users”) at the index visit. The three-month window prior to the index date addresses this to some extent, but to improve the chances of detecting prevalent use, we identified a subset of patients who had a routine primary care visit in the MGB system in the past year, who were thus less likely to have received an undocumented antidepressant from an outside provider. We conducted 200 chart reviews to assess the frequency of prevalent users among both the full cohort and the subset of patients with primary care visits (the “PCP subset”). The analyses described below were performed in both sets of patients.

### Variables for analysis

To determine variables to be included in the analysis, a psychiatrist (YS) searched the literature (3–6) and conducted telephone interviews with five other psychiatrists and three non-psychiatrist physicians. The physicians were first asked open questions regarding what factors they consider when selecting an antidepressant, and then queried more specifically when the initial answers were broad (i.e., drug-drug interaction, etc.) After all information was gathered, a set of variables was derived by including all factors that were mentioned to affect prescribing decisions in either the interviews or the literature. The final list of variables can be broadly categorized as patient demographics, prescription timing information, co-morbidities at the index visit, other medications, and depression-related symptoms.

For co-morbidities, ICD billing codes that occurred before or at the index visit were collected for each patient and mapped to individual diseases, adopting mappings from those either previously validated and published in a peer-reviewed journal (8,9), provided by authoritative sources (10,11), or electronic ICD code databases (12,13) if the prior two sources were not available (Appendix A shows the complete mapping).

Prescriptions between 90 days before the index visit and at the index visit were retrieved for each subject. These medications were then categorized based on generic names regardless of dosing and route of administration. Counts for distinct medications other than antidepressants were then generated for each patient within the 90-day time window. We chose to include the number of medications rather than specific medications because most clinicians reported that this was a bigger driver of decision-making based on differences in broad drug-drug interactions across medication class. However, a count of the number of prescriptions for nonsteroidal anti-inflammatory drugs (NSAIDs) was constructed as a separate variable because these were reported to be specifically associated with abnormal gastrointestinal bleeding with concurrent use of SSRI and SNRI antidepressants (14,15). We also included the calendar year of the index visit because clinical practice may change over time in response to new information, such as new clinical trials or updated guidelines.

### Extracting depression-related symptoms from clinical notes with NLP

We adopted a hierarchical approach to identify and create variables for depression-related symptoms. The hierarchy consisted of four levels, from top to bottom: (1) *categories* of depression-related symptoms (e.g., depressive mood and anhedonia, loss of appetite and body weight, and insomnia), (2) *concepts* within these categories (e.g., for depressive mood and anhedonia: “anhedonia” and “sadness”), (3) *specific terms* used to describe these concepts (e.g., for anhedonia, “anhedonia,” “can’t enjoy anything, “no pleasure”), (4) l*exical derivatives and regular expressions* (a common text format used for computer reading). We initially grouped the concepts of depression-related symptoms into depressive symptom categories corresponding to criteria in the Diagnostic and Statistical Manual of Mental

Disorders-5th edition (DSM-5) (16). We then expanded the search to include terms largely synonymous with these concepts, being fairly liberal in order to make sure we captured the core concept, since many of the terms were derived from the patients’ actual wording; for example, “despondent” was taken as synonymous with “depressed.” The actual variables built into the data matrix were the number of concepts present per category (e.g., category 1 “depressive mood and anhedonia” had a total of 8 concepts, each of which could be indicated by the presence of one of many terms) in the aggregation of notes within the 90-day window prior to and including the index date for each patient. For example, if a patient had at least one term indicating two out of 8 among these concepts, say, “depressed mood” and “melancholia,” he would receive a score of 2 for category 1. Appendix B provides further details on the psychopathology hierarchical structure.

Before extracting terms from the clinical notes, a pre-processing step removed sentence segments containing words that implied negations (e.g., not, no) while sparing double negations and sentence components separated by “but”, “however”, “although”, and “nevertheless” that would indicate a reverse of sentiment. Sentence parsing was done using the spaCy (https://spacy.io/) (17) package for Python (version 3.7), which applied the customized sentence segmentation rules mentioned above. We then applied regular expression matching to detect the presence of any terms for depression-related symptoms within the notes using the Python Re module. Using these matching results, we constructed the depression-related symptom variables in accordance with the hierarchical mapping previously described. To avoid false positive matching, regular expressions of shorter length were protected by word boundary detection (i.e., match only when white spaces or punctuation were present around the string) to prevent them from being matched as a substring of a longer word; for example, “cry” would not be matched if the string only appeared in “cryptococcus.”

## Statistical analysis

### (a) Descriptive analysis of variable value distribution

For each categorical variable, we calculated the proportion of subjects in each category. For continuous variables, we calculated mean, standard deviation, and range. All calculations were performed for the full sample and stratified by index antidepressant category.

### (b) Statistical modeling of initial antidepressant choice

To model the choice of the antidepressant initiated, we performed a multivariate multinomial regression with antidepressant class as the outcome and with SSRIs, the most commonly prescribed class, as the reference category. We included the main effect of each of the 64 variables as predictors in the model, without considering any interactions. We performed the modeling in R version 3.5.2 with the package “mnlogit,” which allows efficient inference using the Newton-Raphson method. We estimated the odds ratio and 95% CI for each variable and for each contrast of treatment with SSRIs (i.e., bupropion, mirtazapine, and SNRI versus SSRI). Likelihood ratio tests were done globally for each variable comparing the full model with the model leaving out the variable of interest with a chi-squared test. This procedure was recurrently applied to generate p-values for each variable. We report both nominal and FDR-corrected p-values.

### (c) Sensitivity analysis

For sensitivity analysis, we identified a subset of patients with routine primary care visits in the MGB system (the “PCP” subset), for whom we were more likely to capture (and thus be able to exclude) prevalent users. To do this, we identified patients with routine health services or annual check-up visits within one year prior to the index date. We identified such patients using the presence of either of the following: (1) at least one of the following Current Procedural Terminology (CPT) codes for these services recorded: 99201, 99202, 99203, 99204, 99205, 99211, 99212, 99213, 99214, 99215; or (2) “reason for visit” recorded as “check-up” in the encounter metadata.

## RESULTS

Our initial selection criteria yielded 111,571 patients from the RPDR. After applying our exclusion criteria, a total of 47,107 patients were retained for the main analysis of the study (“the full sample”). Figure 1 displays the detailed exclusion steps and the resulting patient counts at each step. Among the 47,107 patients, 22,848 had an MGB primary care visit in the year prior to the index date (the “PCP subset”). Table 1.1 presents the descriptive results for the full sample. The majority of patients were first prescribed an SSRI (n = 34,709, 73.7%), followed by bupropion (n = 5,969, 12.7%), SNRI (n = 4,924, 10.4%), and mirtazapine (n = 1,505, 3.2%). The study sample was predominantly female (67%), consistent with the known sex distribution of depression (18). Approximately 77% of the patients were identified as Caucasian. Common comorbid diagnostic codes included anxiety-related diagnoses (34.8%), primary hypertension (29.5%), and any malignancy (26.5%), including past malignancies. The pattern of first prescription class changed over time, as can be seen in Figure 2.

**Table 1:**
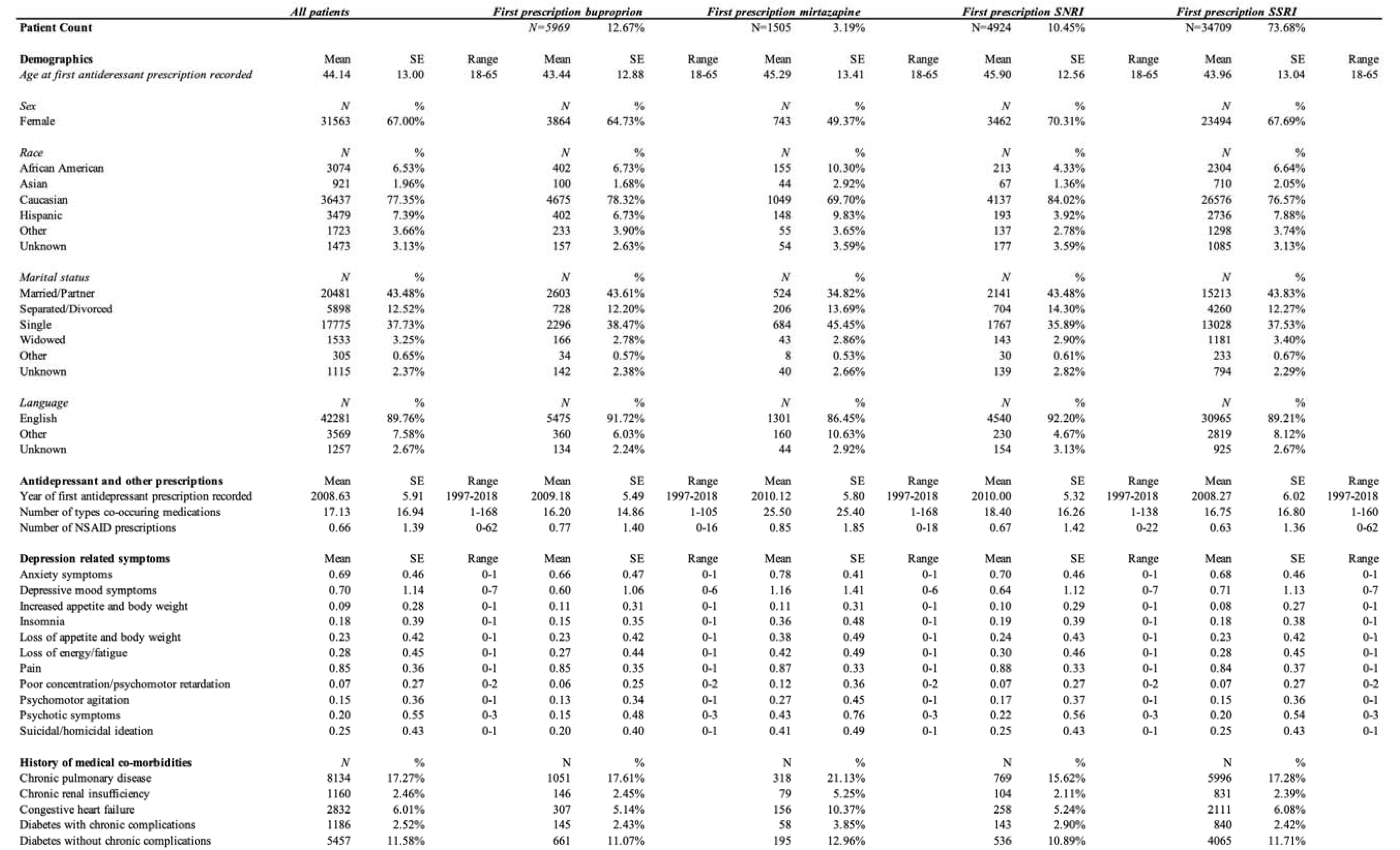
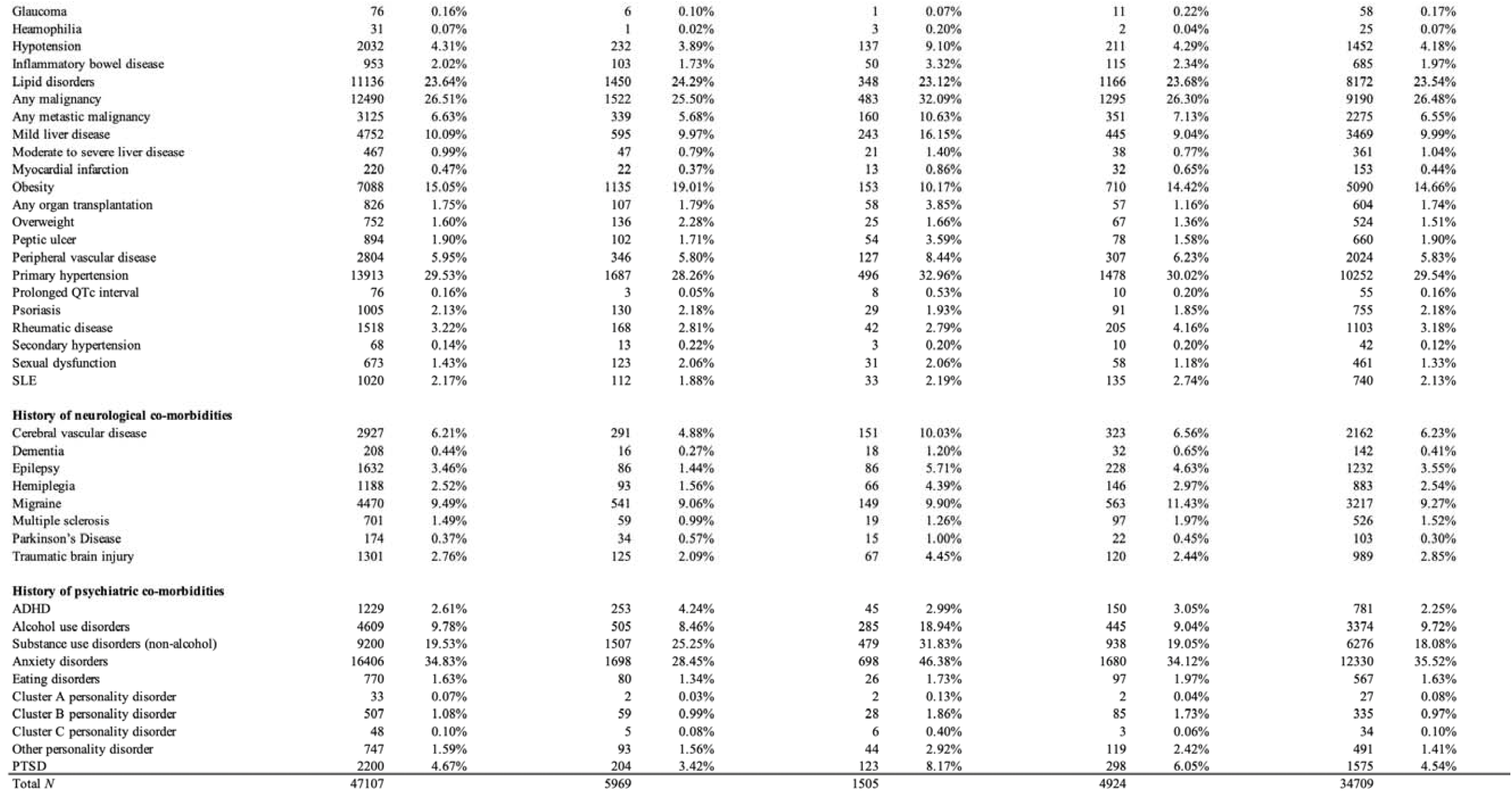
Distribution of patient characteristics (full data set), N=47,107.

**Figure 1:**
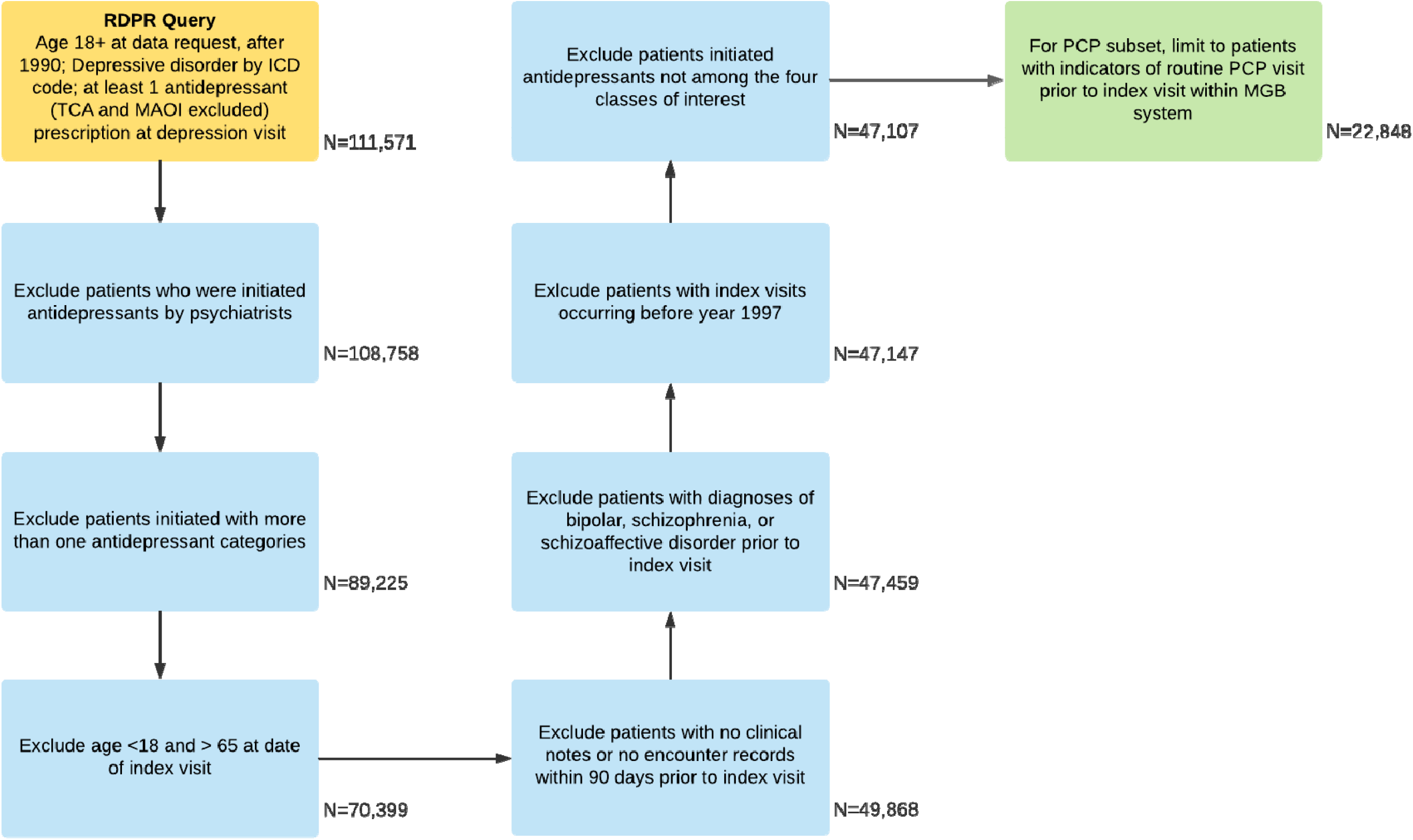
Count of first prescription of antidepressant for depressant per category by year

**Figure 2:**
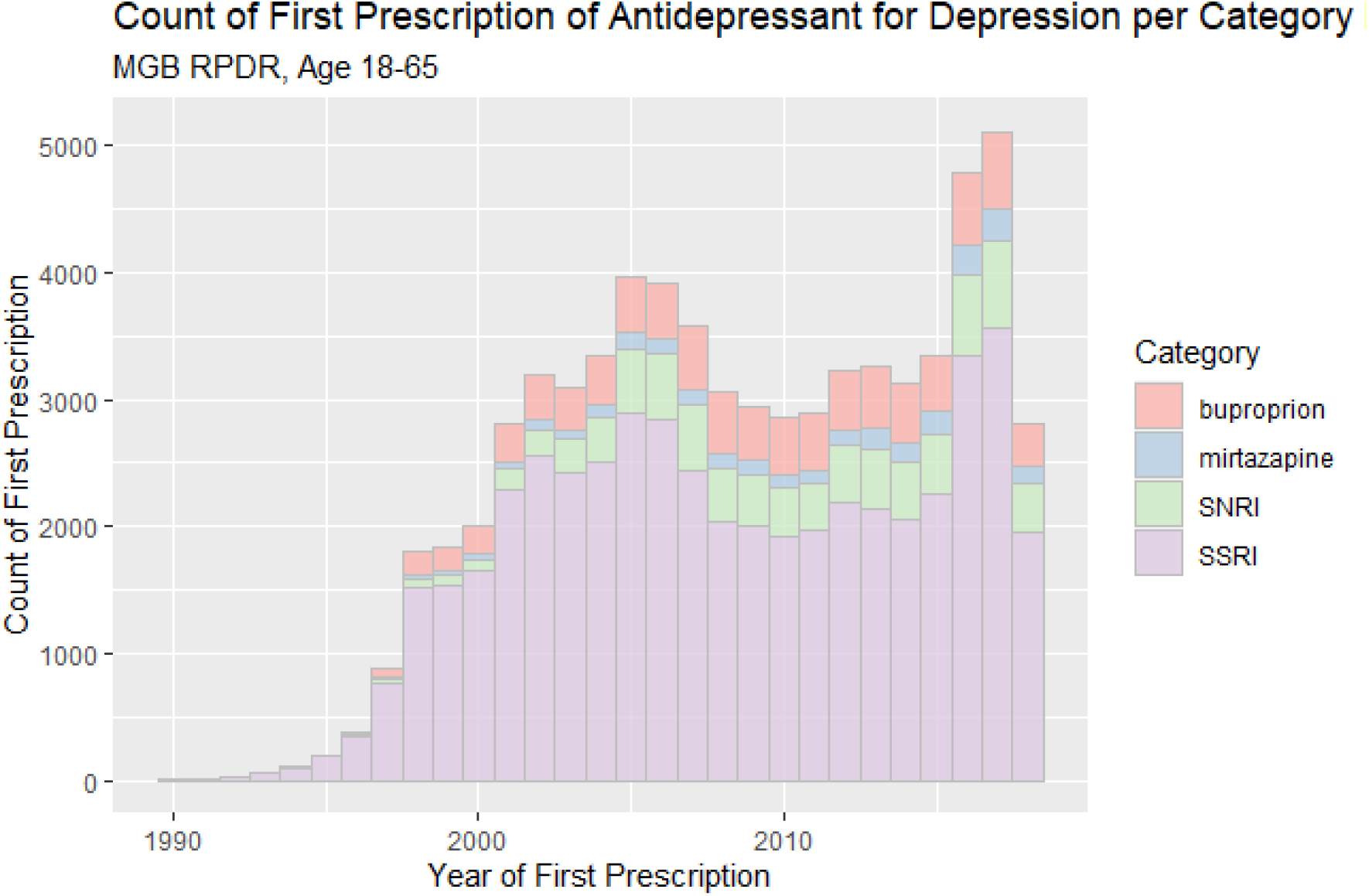
Count of first prescription of antidepressant for depression by category per year, age 18-65

Based on chart reviews of 200 randomly sampled patients, we estimated that approximately 29% of patients in the full sample were not new users. Applying the method above for identifying the PCP subset to the same 200 patients reduced the proportion of non-new users by about one-third--to 19%. The distribution of sociodemographic and clinical features was largely similar between the full sample and the PCP subset (results not shown).

There were statistically significant but small differences in age across the treatment groups, e.g., the SNRI and mirtazapine groups were slightly older (mean ages 45.90 and 45.29, respectively, compared to 43.96 for SSRI). Patients in the bupropion group were more likely to be male (50% vs. 33% overall) and obese (19% vs. 15% overall). The mirtazapine group had higher proportions of co-morbidities (e.g., congestive heart failure, primary hypertension, metastatic malignancy, and problems with sleep), as well as more concomitant medications. Both the bupropion group and the mirtazapine group had higher proportions of co-morbid substance use disorder. The characteristics of the patients in the SSRI and SNRI groups were more similar to one another than to those of the two other groups.

Table 2 shows the association between all selected variables and the choice of antidepressant class in the full sample, and Appendix C for the PCP subset. Figure 3 plots the odds ratios and confidence intervals for variables that were globally significant (i.e., significant for the omnibus test for the variable) in the full sample, (see Appendix D for the PCP subset). Appendix E shows the same plot for all variables in the full sample, and Appendix F for the PCP subset. For the full sample, 36 of 64 candidate predictors identified by literature review and clinical expert consultation were associated with antidepressant class selection. All binary variables among the 36 that were significant had at least one OR of either > 1.1 or < 0.9 for one of the three contrasts (i.e., initiated one of the other three antidepressant categories versus SSRI) derived by multinominal regression). Age, year of prescription, total number of other medications, and number of NSAID prescriptions all showed significant association with treatment selection (all FDR-corrected p < 0.001). Among all psychiatric comorbidities considered, only eating disorders and cluster A/B/C personality disorders were not associated with initial antidepressant selection. Other psychiatric disorders, including attention-deficit/hyperactivity disorder, alcohol use disorders, other substance use disorders, anxiety disorders, post-traumatic stress disorder, and other personality disorders, all showed strong signals of association (FDR-corrected p = 0.002 for other personality disorders; p <0.001 for all other diagnoses).

**Table 2:** Modeled propensity odds ratio and 95% confidence intervals, full data set.

|  | *Bupropion vs SSRI* |  |  | *Mirtazapine vs SSRI* |  |  | *SNRI vsSSRI* |  |  |  |  |
| --- | --- | --- | --- | --- | --- | --- | --- | --- | --- | --- | --- |
|  | Odds ratio | 95% CI |  | Odds ratio | 95% CI |  | Odds ratio | 95% CI |  | Raw global *p*-value | Corrected global *p*-value |
| **Demographics** |  |  |  |  |  |  |  |  |  |  |  |
| Age at antidepressant prescription (per year increase) | 0.997 | 0.995 | 1.000 | 1.015 | 1.010 | 1.020 | 1.015 | 1.012 | 1.018 | >0.001 | >0.001 *** |
| Sex: Male | 1.137 | 1.067 | 1.212 | 1.768 | 1.574 | 1.985 | 0.826 | 0.769 | 0.887 | >0.001 | >0.001 *** |
| Race: African American | 0.998 | 0.889 | 1.120 | 1.568 | 1.301 | 1.889 | 0.597 | 0.514 | 0.692 |  |  |
| Race: Asian | 0.873 | 0.703 | 1.085 | 1.403 | 1.011 | 1.947 | 0.630 | 0.487 | 0.814 |  |  |
| Race: Hispanic | 0.941 | 0.829 | 1.068 | 1.260 | 1.015 | 1.563 | 0.549 | 0.465 | 0.650 |  |  |
| Race: Other | 1.106 | 0.946 | 1.292 | 0.953 | 0.705 | 1.288 | 0.746 | 0.616 | 0.905 |  |  |
| Race: Unknown | 0.887 | 0.743 | 1.059 | 1.168 | 0.866 | 1.573 | 1.045 | 0.880 | 1.240 | >0.001 | >0.001 *** |
| Marital status: Married/Partner | 1.020 | 0.953 | 1.091 | 0.708 | 0.621 | 0.808 | 0.908 | 0.843 | 0.978 |  |  |
| Marital status: Separated/Divorced | 1.059 | 0.962 | 1.167 | 0.962 | 0.811 | 1.142 | 1.113 | 1.007 | 1.231 |  |  |
| Marital status: Widowed | 0.946 | 0.793 | 1.130 | 0.821 | 0.590 | 1.144 | 0.813 | 0.673 | 0.982 | >0.001 | >0.001 *** |
| Marital status: Other | 0.889 | 0.615 | 1.284 | 0.711 | 0.343 | 1.477 | 0.855 | 0.580 | 1.260 |  |  |
| Marital status: Unknown | 1.131 | 0.934 | 1.370 | 1.050 | 0.744 | 1.482 | 1.136 | 0.934 | 1.382 |  |  |
| Language: Other | 0.772 | 0.675 | 0.882 | 1.284 | 1.041 | 1.584 | 0.726 | 0.619 | 0.852 |  |  |
| Language: Unknown | 0.893 | 0.740 | 1.078 | 1.040 | 0.756 | 1.430 | 1.119 | 0.936 | 1.338 | >0.001 | >0.001 *** |
| **Antidepressant and other prescriptions** |  |  |  |  |  |  |  |  |  |  |  |
| Year of first prescription (per year increase) | 1.039 | 1.034 | 1.045 | 1.034 | 1.024 | 1.045 | 1.060 | 1.054 | 1.066 | >0.001 | >0.001 *** |
| Number of types of co-occurring medications (per number increase) | 0.996 | 0.994 | 0.998 | 1.016 | 1.013 | 1.019 | 1.003 | 1.001 | 1.006 | >0.001 | >0.001 *** |
| Number of NSAID prescriptions (per number increase) | 1.076 | 1.055 | 1.097 | 1.012 | 0.978 | 1.046 | 0.998 | 0.975 | 1.022 | >0.001 | >0.001 *** |
| **Depression related symptoms** |  |  |  |  |  |  |  |  |  |  |  |
| Anxiety symptoms | 0.947 | 0.888 | 1.010 | 0.997 | 0.867 | 1.145 | 0.992 | 0.923 | 1.066 | 0.429 | 0.458 |
| Depressive mood and anhedonia (per level increase) | 0.937 | 0.904 | 0.970 | 1.048 | 0.993 | 1.106 | 0.895 | 0.862 | 0.929 | >0.001 | >0.001 *** |
| Increased appetite and body weight | 1.429 | 1.298 | 1.572 | 0.981 | 0.819 | 1.174 | 1.178 | 1.057 | 1.313 | >0.001 | >0.001 *** |
| Insomnia | 0.858 | 0.789 | 0.933 | 1.582 | 1.394 | 1.795 | 1.063 | 0.977 | 1.157 | >0.001 | >0.001 *** |
| Loss of appetite and body weight | 1.018 | 0.947 | 1.094 | 1.327 | 1.173 | 1.501 | 1.004 | 0.929 | 1.085 | >0.001 | >0.001 *** |
| Loss of energy/fatigue | 0.971 | 0.907 | 1.040 | 1.125 | 0.995 | 1.272 | 1.019 | 0.948 | 1.096 | 0.192 | 0.233 |
| Pain | 1.031 | 0.949 | 1.119 | 0.920 | 0.780 | 1.084 | 1.216 | 1.106 | 1.338 | >0.001 | 0.001 ** |
| Poor concentration/psychomotor retardation (per level increase) | 1.095 | 0.969 | 1.237 | 0.823 | 0.691 | 0.979 | 1.025 | 0.905 | 1.161 | 0.048 | 0.082 |
| Psychomotor agitation | 0.929 | 0.850 | 1.015 | 1.079 | 0.941 | 1.236 | 1.009 | 0.922 | 1.103 | 0.234 | 0.265 |
| Psychotic symptoms (per level increase) | 0.948 | 0.884 | 1.017 | 1.114 | 1.019 | 1.219 | 1.096 | 1.025 | 1.171 | 0.001 | 0.003 ** |
| Suicidal tendency | 0.760 | 0.696 | 0.829 | 1.296 | 1.121 | 1.498 | 1.068 | 0.976 | 1.169 | >0.001 | >0.001 *** |
| **History of medical co-morbidities** |  |  |  |  |  |  |  |  |  |  |  |
| Chronic pulmonary disease | 0.995 | 0.922 | 1.073 | 1.092 | 0.953 | 1.251 | 0.869 | 0.798 | 0.946 | 0.005 | 0.011 * |
| Chronic renal insufficiency | 1.032 | 0.842 | 1.265 | 1.131 | 0.850 | 1.503 | 0.807 | 0.641 | 1.016 | 0.191 | 0.233 |
| Congestive heart failure | 0.856 | 0.747 | 0.980 | 1.088 | 0.888 | 1.334 | 0.859 | 0.742 | 0.994 | 0.021 | 0.038 * |
| Diabetes with chronic complications | 1.084 | 0.883 | 1.331 | 1.147 | 0.824 | 1.597 | 1.401 | 1.137 | 1.727 | 0.019 | 0.034 * |
| Diabetes without chronic complications | 0.916 | 0.827 | 1.016 | 0.865 | 0.716 | 1.045 | 0.927 | 0.828 | 1.039 | 0.123 | 0.173 |
| Glaucoma | 0.632 | 0.269 | 1.484 | 0.222 | 0.030 | 1.658 | 1.026 | 0.530 | 1.985 | 0.203 | 0.241 |
| Heamophilia | 0.222 | 0.030 | 1.656 | 1.787 | 0.506 | 6.308 | 0.651 | 0.151 | 2.803 | 0.193 | 0.233 |
| Hypotension | 1.027 | 0.883 | 1.194 | 1.127 | 0.916 | 1.387 | 0.884 | 0.756 | 1.035 | 0.236 | 0.265 |
| Inflammatory bowel disease | 0.873 | 0.705 | 1.080 | 1.466 | 1.080 | 1.989 | 1.182 | 0.964 | 1.449 | 0.017 | 0.032 * |
| Lipid disorders | 1.029 | 0.954 | 1.110 | 0.901 | 0.780 | 1.041 | 0.958 | 0.883 | 1.040 | 0.285 | 0.315 |
| Any malignancy | 0.972 | 0.905 | 1.044 | 1.009 | 0.881 | 1.156 | 0.902 | 0.834 | 0.975 | 0.070 | 0.111 |
| Any metastatic malignancy | 0.901 | 0.790 | 1.028 | 1.332 | 1.087 | 1.633 | 0.990 | 0.866 | 1.130 | 0.014 | 0.029 * |
| Mild liver disease | 1.018 | 0.921 | 1.126 | 1.179 | 1.005 | 1.383 | 0.932 | 0.832 | 1.043 | 0.104 | 0.152 |
| Moderate to severe liver disease | 0.798 | 0.578 | 1.102 | 0.522 | 0.324 | 0.842 | 0.802 | 0.564 | 1.142 | 0.016 | 0.032 * |
| History of myocardial infarction | 0.707 | 0.444 | 1.124 | 0.706 | 0.377 | 1.320 | 0.928 | 0.624 | 1.380 | 0.349 | 0.378 |
| Obesity | 1.343 | 1.240 | 1.455 | 0.607 | 0.504 | 0.731 | 0.983 | 0.895 | 1.080 | >0.001 | >0.001 *** |
| Any organ transplantation | 1.329 | 1.059 | 1.667 | 1.312 | 0.956 | 1.801 | 0.775 | 0.581 | 1.035 | 0.005 | 0.012 * |
| Overweight | 1.151 | 0.942 | 1.406 | 1.047 | 0.684 | 1.605 | 0.807 | 0.620 | 1.050 | 0.145 | 0.189 |
| Peptic ulcer | 0.957 | 0.770 | 1.188 | 1.447 | 1.072 | 1.953 | 0.926 | 0.727 | 1.180 | 0.097 | 0.148 |
| Peripheral vascular disease | 1.041 | 0.915 | 1.186 | 0.970 | 0.784 | 1.201 | 1.010 | 0.882 | 1.157 | 0.922 | 0.922 |
| Primary hypertension | 0.937 | 0.870 | 1.009 | 0.866 | 0.758 | 0.990 | 0.999 | 0.924 | 1.081 | 0.071 | 0.111 |
| Prolonged QTc interval | 0.335 | 0.104 | 1.079 | 1.340 | 0.614 | 2.926 | 0.842 | 0.424 | 1.671 | 0.130 | 0.173 |
| Psoriasis | 0.950 | 0.783 | 1.152 | 0.865 | 0.589 | 1.270 | 0.800 | 0.640 | 1.001 | 0.218 | 0.254 |

**Table 2: Modeled propensity odds ratio and 95% confidence intervals, full data set (Continued)**
|  |  |  |  |  |  |  |  |  |  |  |  |
| --- | --- | --- | --- | --- | --- | --- | --- | --- | --- | --- | --- |
| Rheumatic disease | 0.952 | 0.781 | 1.161 | 0.906 | 0.623 | 1.318 | 1.360 | 1.132 | 1.635 | 0.008 | 0.017 * |
| Secondary hypertension | 1.816 | 0.953 | 3.464 | 0.823 | 0.243 | 2.782 | 1.797 | 0.885 | 3.648 | 0.174 | 0.222 |
| SLE | 0.911 | 0.715 | 1.160 | 0.979 | 0.639 | 1.499 | 0.974 | 0.779 | 1.218 | 0.038 | 0.911 |
| Sexual dysfunction | 1.378 | 1.115 | 1.703 | 1.038 | 0.704 | 1.530 | 1.035 | 0.780 | 1.374 | 0.897 | 0.066 |
| **History of neurological co-morbidities** |  |  |  |  |  |  |  |  |  |  |  |
| Cerebral vascular disease | 0.879 | 0.764 | 1.012 | 1.151 | 0.935 | 1.416 | 0.956 | 0.834 | 1.097 | 0.127 | 0.173 |
| Dementia | 0.832 | 0.491 | 1.410 | 1.504 | 0.891 | 2.541 | 1.436 | 0.965 | 2.137 | 0.128 | 0.173 |
| Epilepsy | 0.411 | 0.328 | 0.514 | 1.053 | 0.828 | 1.340 | 1.115 | 0.959 | 1.296 | >0.001 | >0.001 *** |
| Hemiplegia | 0.692 | 0.553 | 0.867 | 1.142 | 0.862 | 1.512 | 1.123 | 0.930 | 1.357 | 0.002 | 0.005 ** |
| Migraine | 1.026 | 0.929 | 1.134 | 1.227 | 1.021 | 1.474 | 1.333 | 1.206 | 1.473 | >0.001 | >0.001 *** |
| Multiple sclerosis | 0.689 | 0.524 | 0.906 | 0.999 | 0.623 | 1.601 | 1.241 | 0.992 | 1.551 | 0.005 | 0.011 * |
| Parkinsons disease | 2.112 | 1.419 | 3.143 | 2.960 | 1.668 | 5.254 | 1.259 | 0.787 | 2.012 | >0.001 | >0.001 *** |
| History of traumatic brain injury | 0.775 | 0.638 | 0.941 | 0.980 | 0.746 | 1.286 | 0.811 | 0.665 | 0.990 | 0.017 | 0.032 * |
| **History of psychiatric co-morbidities** |  |  |  |  |  |  |  |  |  |  |  |
| ADHD | 1.815 | 1.560 | 2.110 | 1.074 | 0.782 | 1.475 | 1.229 | 1.024 | 1.476 | >0.001 | >0.001 *** |
| Alcohol use disorders | 0.657 | 0.586 | 0.736 | 1.014 | 0.857 | 1.200 | 0.812 | 0.719 | 0.918 | >0.001 | >0.001 *** |
| Substance use disorders (non-alcohol) | 2.018 | 1.871 | 2.175 | 1.474 | 1.282 | 1.694 | 1.239 | 1.134 | 1.355 | >0.001 | >0.001 *** |
| Any anxiety disorders | 0.677 | 0.633 | 0.724 | 1.206 | 1.073 | 1.355 | 0.832 | 0.776 | 0.893 | >0.001 | >0.001 *** |
| Any eating disorders | 0.817 | 0.641 | 1.041 | 1.246 | 0.826 | 1.879 | 1.160 | 0.925 | 1.454 | 0.103 | 0.152 |
| Cluster A personality disorder | 0.494 | 0.115 | 2.125 | 1.135 | 0.264 | 4.882 | 0.505 | 0.118 | 2.168 | 0.574 | 0.603 |
| Cluster B personality disorder | 1.069 | 0.743 | 1.538 | 1.056 | 0.637 | 1.748 | 1.249 | 0.897 | 1.738 | 0.630 | 0.650 |
| Cluster C personality disorder | 0.968 | 0.371 | 2.521 | 3.893 | 1.588 | 9.542 | 0.657 | 0.199 | 2.165 | 0.055 | 0.091 |
| Other personality disorders | 1.355 | 1.011 | 1.817 | 1.492 | 0.991 | 2.246 | 1.718 | 1.299 | 2.271 | 0.001 | 0.002 ** |
| PTSD | 0.870 | 0.742 | 1.020 | 1.169 | 0.946 | 1.443 | 1.405 | 1.221 | 1.617 | >0.001 | >0.001 *** |

\*: $p < 0.05$ ; \*\*: $p < 0.01$ ; \*\*\*: $p < 0.001$

**Figure 3:**
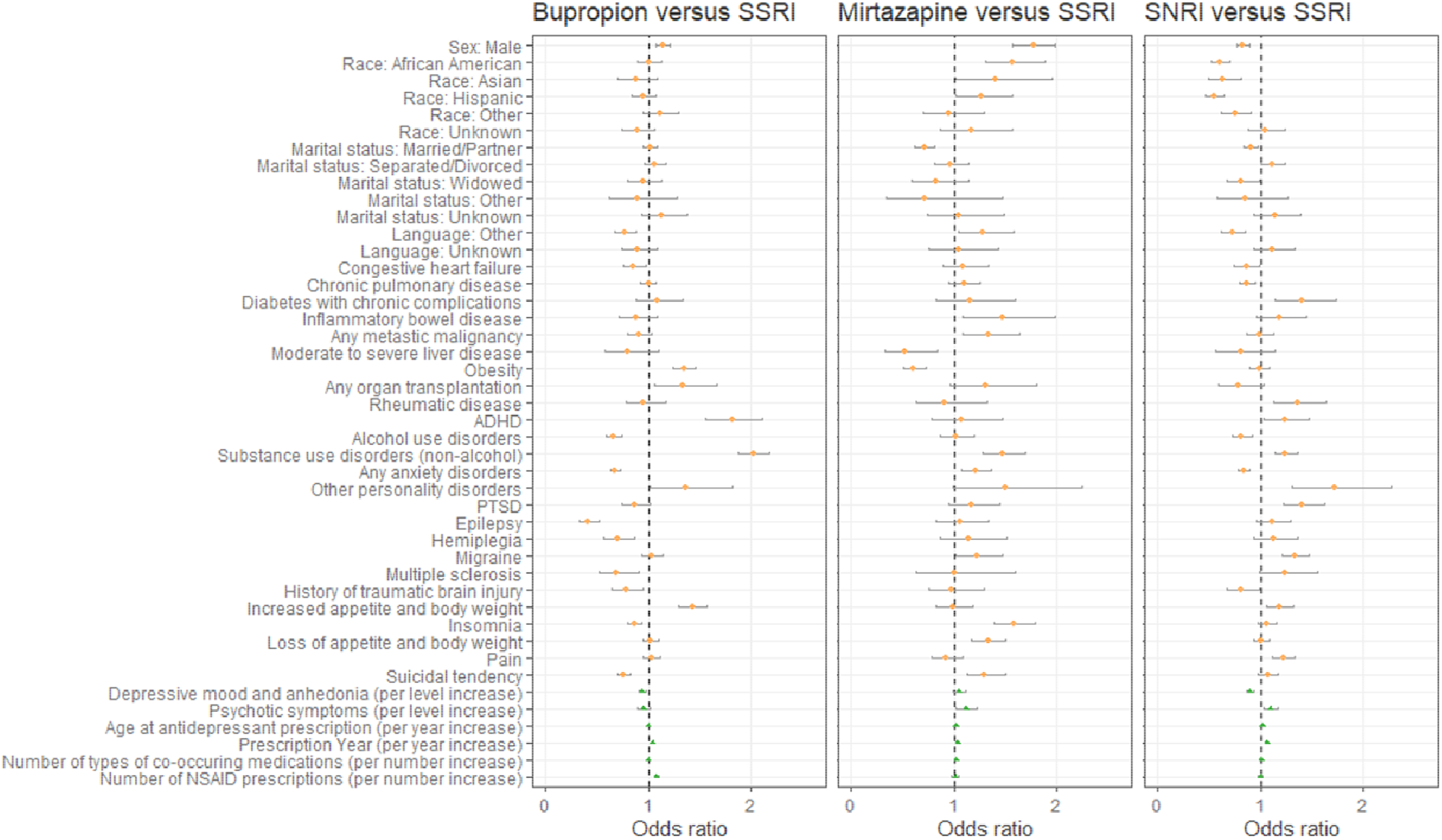
Forest plot for ORs and CIs of globally significant variables for multinominal regression model on antidepressant category propensity, full data set. *Parkinson’s disease not shown for illustration purpose due to very wide confidence interval

With the exception of cerebrovascular disease and stroke, all examined neurological comorbidities were associated with antidepressant selection. Notably, bupropion was less commonly prescribed to patients with neurological disorders, such as epilepsy, hemiplegia, multiple sclerosis, traumatic brain injury, and cerebral vascular disease, while bupropion was more commonly prescribed to patients with Parkinson’s disease. Mirtazapine and SNRIs were more commonly prescribed to patients with migraine and hemiplegia, while SNRIs were less commonly prescribed to patients with cardiovascular disease.

As for general medical comorbidities, bupropion was less commonly prescribed to patients with congestive heart failure, and was more commonly given to patients with obesity, organ transplantation, and sexual dysfunction. Mirtazapine was less frequently prescribed for patients with obesity, moderate to severe liver disease, and primary hypertension, and more frequently prescribed for patients with inflammatory bowel disease, mild liver disease, metastatic malignancy, organ transplantation, and peptic ulcer, compared to SSRIs, with all other factors controlled. SNRIs were more frequently prescribed to patients with diabetes with chronic complications and less frequently to patients with congestive heart failure and chronic pulmonary disease.

As can be seen in Appendix C, Appendix D, and Appendix F, despite some loss of power, the general pattern of findings was very similar in the PCP subset.

## DISCUSSION

Using real-world EHR data to study the initial prescribing choices for depression among non-psychiatrists, the current study detected strong signals for many factors consistent with currently recommended psychiatric practice. For example, pain and loss of appetite showed strong associations with SNRIs (which have indications in pain management) and mirtazapine (which is known to increase appetite). Applying NLP to unstructured EHR enabled the current study to capture depression-related clinical characteristics, even in non-psychiatric settings where relevant records might be sparse. Recent studies have also looked into antidepressant prescriptions in non-psychiatric settings, such as off-label use of antidepressants (19), as well as modeling the overall likelihood of any treatment initiation for people with depression in primary care settings (20). One study (21) performed semi-structured interviews with 28 general practitioners regarding the factors they would consider when prescribing an antidepressant. However, none of these studies quantitatively analyzed the association between detailed clinical characteristics and the choice of antidepressant with depression as the indication, as we have done here.

It is of particular interest that most of the neurological disorders studied showed significant correlations with the choice of antidepressant. The inverse association of bupropion prescription with epilepsy and other neurological disorders, such as traumatic brain injury (22), multiple sclerosis (23), and hemiplegia (most commonly caused by stroke) (24) could be explained by prescribing physicians taking into account bupropion’s lowering of the seizure threshold. On the other hand, the association of mirtazapine prescription with comorbid migraine was unexpected, since there are no obvious direct pharmacological or clinical considerations that would support this association.

Our results should be interpreted in the context of several limitations. First, real-world EHR data are inherently limited by the presence of missing data. In our case, data were present only when the patient received care within the MGB system and only for observations documented in the EHR. As mentioned previously, another limitation of our study is the difficulty of discerning new from prevalent users. This is also a problem of missing data in the sense that we might be missing antidepressant prescription information prior to the index visit, particularly for patients who obtain initial prescriptions from community psychiatrists or other sources. With prevalent users, the characteristics collected in the window before the index visit might not reflect the patient’s status in the period before actual treatment initiation, which may have occurred long before the index visit date. It is also possible that some of these characteristics, could have been *caused b*y the prevalent drug (e.g., loss of appetite and weight loss can be caused by bupropion) and thus adjusting for them could introduce bias. To address this, our sensitivity analysis looked at patients with PCPs within the MGB system, where the proportion of prevalent users as judged by physician chart review was somewhat lower (29% vs. 19%); these analyses yielded results similar to the main analysis. That said, we acknowledge these problems could be further mitigated by obtaining more complete information regarding treatment and health histories (e.g., by mining medication lists beyond prescriptions or clinical notes for evidence of medications patients are taking from outside sources, by linking EHR data to insurance claims data that capture encounters and treatment beyond a single healthcare system, or by linking specifically to part D claims).

Second, phenotyping depression with ICD codes alone may result in low sensitivity and specificity. This can be remedied to some extent by applying phenotyping algorithms that can be more accurate than ICD codes alone (25–27). We elected not to use such algorithms here because we were interested in prescribing practices when the non-psychiatrist believes that they are treating depression, irrespective of the accuracy of such a diagnosis.

A third limitation is that we did not consider non-pharmacological therapies in the study. Part of the reason for not considering psychotherapies is that a substantial fraction of psychotherapy may occur outside the MGB system. Thus, we were not able to address whether such treatments had been used instead of pharmacologic treatments, before pharmacologic treatments, or concurrent with them. That said, it has been documented that specialized mental health services are less likely to be recommended in primary care settings (28), and this likely applies to other non-psychiatrist clinicians as well.

Fourth, we did not examine antidepressant dosing. The dosing of the medications may affect the observed choice for initiation for drugs for indications other than depression (e.g., low dose mirtazapine for insomnia). Since insomnia is itself a symptom of depression and the two frequently co-occur, without dosing information it would be difficult to discern the actual treatment target in such cases. One may argue that factors considered by the physician upon prescription may be different for the two indications; alternatively, use of mirtazapine to treat primary insomnia might upwardly bias an apparent association between this symptom and mirtazapine selection. In addition, primary care physicians may use low doses but still be intending to treat depression—and in fact this is likely given the presence of a depression code for the visit at drug initiation. Finally, because our data are observational, we cannot be sure that factors associated with prescription were the cause of prescription choices.

In conclusion, our study investigated factors associated with first prescription choice of antidepressants by non-psychiatrists using large-scale EHR data, incorporating both structured and unstructured features. To our knowledge, this is the first study to quantitatively demonstrate that factors affecting the choice of first antidepressant prescription by non-psychiatrists are generally consistent with those recommended or commonly used in psychiatric practice. Future research may benefit from efforts to improve data completeness and cleanliness, phenotyping accuracy, and natural language processing techniques.

## Supporting information

STROBE checklist

## Data Availability

The electronic health records data used in this study are not available either to the public or per request for privacy protection.

## ACKNOWLEDGEMENTS

The authors thank the MGB RPDR team for providing the EHR data in an analyzable format.

## CONFLICT OF INTEREST

JWS is a member of the Leon Levy Foundation Neuroscience Advisory Board and received an honorarium for an internal seminar at Biogen, Inc. He is PI of a collaborative study of the genetics of depression and bipolar disorder sponsored by 23andMe for which 23andMe provides analysis time as in-kind support but no payments.

## Appendix A ICD codes mapping to identifying pre-existing co-morbid conditions

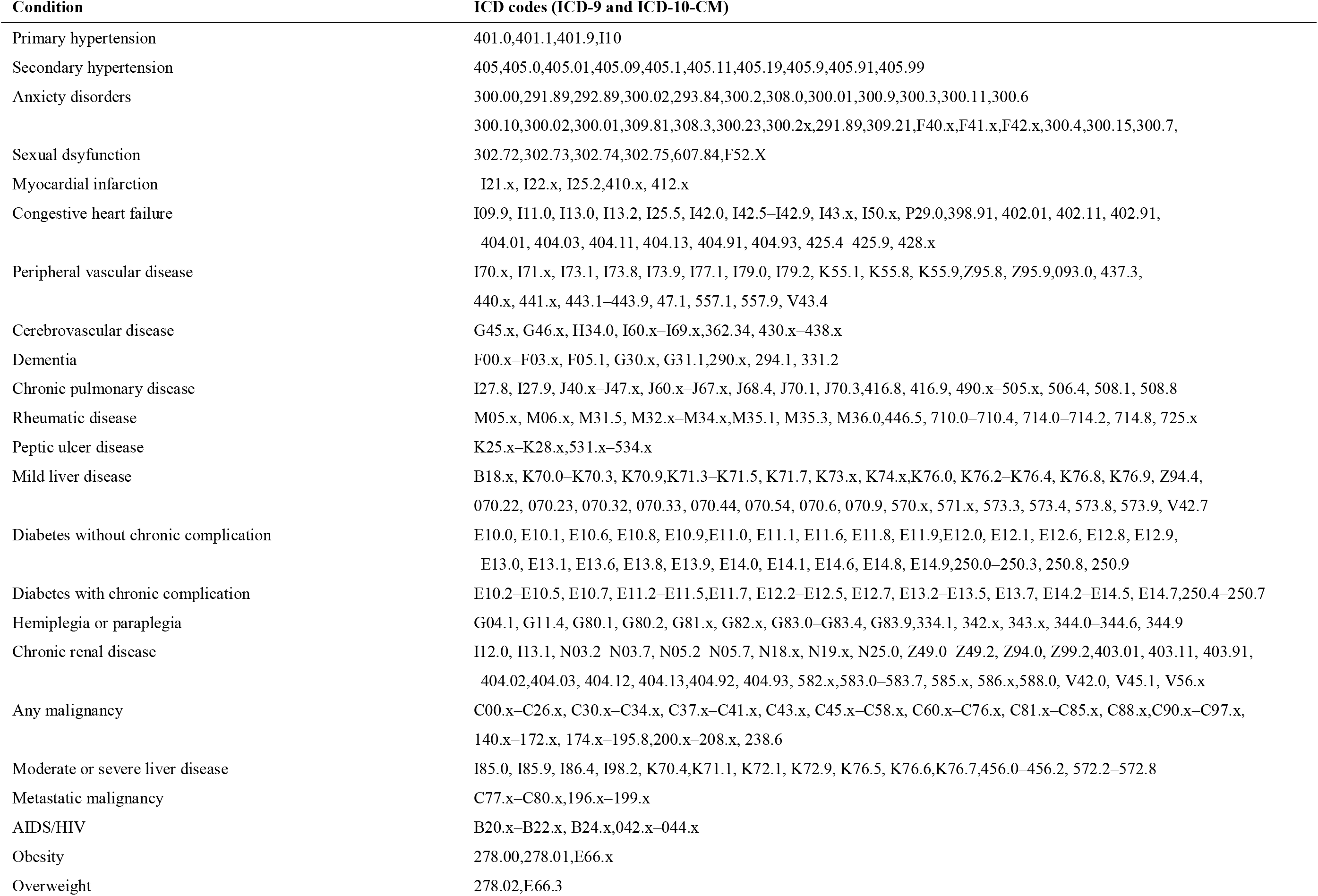
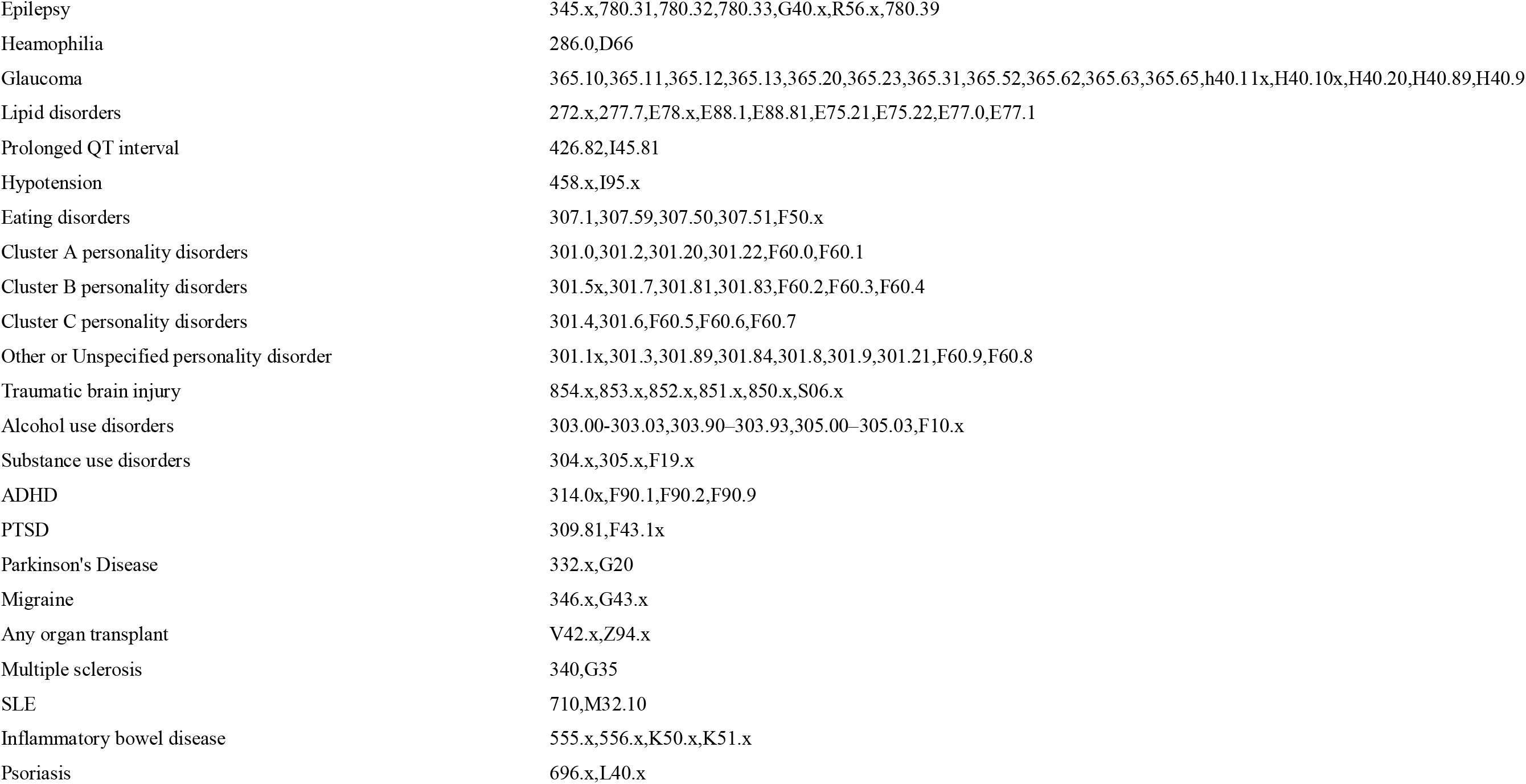

Appendix A: ICD codes mapping to identifying pre-existing co-morbid conditions
| Condition | ICD codes (ICD-9 and ICD-10-CM) |
| --- | --- |
| Primary hypertension | 401.0,401.1,401.9,I10 |
| Secondary hypertension | 405,405.0,405.01,405.09,405.1,405.11,405.19,405.9,405.91,405.99 |
| Anxiety disorders | 300.00,291.89,292.89,300.02,293.84,300.2,308.0,300.01,300.9,300.3,300.11,300.6<br>300.10,300.02,300.01,309.81,308.3,300.23,300.2x,291.89,309.21,F40.x,F41.x,F42.x,300.4,300.15,300.7, |
| Sexual dysfunction | 302.72,302.73,302.74,302.75,607.84,F52.X |
| Myocardial infarction | I21.x, I22.x, I25.2,410.x, 412.x |
| Congestive heart failure | I09.9, I11.0, I13.0, I13.2, I25.5, I42.0, I42.5–I42.9, I43.x, I50.x, P29.0,398.91, 402.01, 402.11, 402.91,<br>404.01, 404.03, 404.11, 404.13, 404.91, 404.93, 425.4–425.9, 428.x |
| Peripheral vascular disease | I70.x, I71.x, I73.1, I73.8, I73.9, I77.1, I79.0, I79.2, K55.1, K55.8, K55.9,Z95.8, Z95.9,093.0, 437.3,<br>440.x, 441.x, 443.1–443.9, 47.1, 557.1, 557.9, V43.4 |
| Cerebrovascular disease | G45.x, G46.x, H34.0, I60.x–I69.x,362.34, 430.x–438.x |
| Dementia | F00.x–F03.x, F05.1, G30.x, G31.1,290.x, 294.1, 331.2 |
| Chronic pulmonary disease | I27.8, I27.9, J40.x–J47.x, J60.x–J67.x, J68.4, J70.1, J70.3,416.8, 416.9, 490.x–505.x, 506.4, 508.1, 508.8 |
| Rheumatic disease | M05.x, M06.x, M31.5, M32.x–M34.x,M35.1, M35.3, M36.0,446.5, 710.0–710.4, 714.0–714.2, 714.8, 725.x |
| Peptic ulcer disease | K25.x–K28.x,531.x–534.x |
| Mild liver disease | B18.x, K70.0–K70.3, K70.9,K71.3–K71.5, K71.7, K73.x, K74.x,K76.0, K76.2–K76.4, K76.8, K76.9, Z94.4,<br>070.22, 070.23, 070.32, 070.33, 070.44, 070.54, 070.6, 070.9, 570.x, 571.x, 573.3, 573.4, 573.8, 573.9, V42.7 |
| Diabetes without chronic complication | E10.0, E10.1, E10.6, E10.8, E10.9,E11.0, E11.1, E11.6, E11.8, E11.9,E12.0, E12.1, E12.6, E12.8, E12.9,<br>E13.0, E13.1, E13.6, E13.8, E13.9, E14.0, E14.1, E14.6, E14.8, E14.9,250.0–250.3, 250.8, 250.9 |
| Diabetes with chronic complication | E10.2–E10.5, E10.7, E11.2–E11.5,E11.7, E12.2–E12.5, E12.7, E13.2–E13.5, E13.7, E14.2–E14.5, E14.7,250.4–250.7 |
| Hemiplegia or paraplegia | G04.1, G11.4, G80.1, G80.2, G81.x, G82.x, G83.0–G83.4, G83.9,334.1, 342.x, 343.x, 344.0–344.6, 344.9 |
| Chronic renal disease | I12.0, I13.1, N03.2–N03.7, N05.2–N05.7, N18.x, N19.x, N25.0, Z49.0–Z49.2, Z94.0, Z99.2,403.01, 403.11, 403.91,<br>404.02,404.03, 404.12, 404.13,404.92, 404.93, 582.x,583.0–583.7, 585.x, 586.x,588.0, V42.0, V45.1, V56.x |
| Any malignancy | C00.x–C26.x, C30.x–C34.x, C37.x–C41.x, C43.x, C45.x–C58.x, C60.x–C76.x, C81.x–C85.x, C88.x,C90.x–C97.x,<br>140.x–172.x, 174.x–195.8,200.x–208.x, 238.6 |
| Moderate or severe liver disease | I85.0, I85.9, I86.4, I98.2, K70.4,K71.1, K72.1, K72.9, K76.5, K76.6,K76.7,456.0–456.2, 572.2–572.8 |
| Metastatic malignancy | C77.x–C80.x,196.x–199.x |
| AIDS/HIV | B20.x–B22.x, B24.x,042.x–044.x |
| Obesity | 278.00,278.01,E66.x |
| Overweight | 278.02,E66.3 |
| Epilepsy | 345.x,780.31,780.32,780.33,G40.x,R56.x,780.39 |
| Heamophilia | 286.0,D66 |
| Glaucoma | 365.10,365.11,365.12,365.13,365.20,365.23,365.31,365.52,365.62,365.63,365.65,h40.11x,H40.10x,H40.20,H40.89,H40.9 |
| Lipid disorders | 272.x,277.7,E78.x,E88.1,E88.81,E75.21,E75.22,E77.0,E77.1 |
| Prolonged QT interval | 426.82,I45.81 |
| Hypotension | 458.x,I95.x |
| Eating disorders | 307.1,307.59,307.50,307.51,F50.x |
| Cluster A personality disorders | 301.0,301.2,301.20,301.22,F60.0,F60.1 |
| Cluster B personality disorders | 301.5x,301.7,301.81,301.83,F60.2,F60.3,F60.4 |
| Cluster C personality disorders | 301.4,301.6,F60.5,F60.6,F60.7 |
| Other or Unspecified personality disorder | 301.1x,301.3,301.89,301.84,301.8,301.9,301.21,F60.9,F60.8 |
| Traumatic brain injury | 854.x,853.x,852.x,851.x,850.x,S06.x |
| Alcohol use disorders | 303.00-303.03,303.90–303.93,305.00–305.03,F10.x |
| Substance use disorders | 304.x,305.x,F19.x |
| ADHD | 314.0x,F90.1,F90.2,F90.9 |
| PTSD | 309.81,F43.1x |
| Parkinson's Disease | 332.x,G20 |
| Migraine | 346.x,G43.x |
| Any organ transplant | V42.x,Z94.x |
| Multiple sclerosis | 340,G35 |
| SLE | 710,M32.10 |
| Inflammatory bowel disease | 555.x,556.x,K50.x,K51.x |
| Psoriasis | 696.x,L40.x |

## Appendix B Concept terms table for depression-related symptoms construction

*This table is constructed starting from the “concept names” (the third column), which are intended to capture the criteria for major depressive episode in DSM-5. Concepts are grouped into categories (the second column), which are given a numeric index (the first column). Each concept is described by one or more terms (the 4th column). Lexical derivatives and their matching regular expression (what the computer reads) are then constructed based on the terms (not shown).

| *Category No.* | *Category Name* | *Concept Name* | *Concept Terms* |
| --- | --- | --- | --- |
| 1 | Depressive mood and anhedonia | Depressed mood<br>Guilty feeling<br>Melancholia<br>Hopelessness<br>Anhedonia/loss of interest<br>Weepiness<br>Pessimism<br>Worthlessness | dejected, depressed, despondent, dispirited, dragged, morose, unhappy<br>guilt<br>melancholia<br>forlorn, helpless, hopeless<br>anhedonia, diminished interest, loss of interest, loss of motivation<br>crying, sad, sorrow, tearful, weep<br>Pessimistic, pessimistic thoughts<br>low self esteem, unworthiness, worthlessness |
| 2 | Poor concentration/psychomotor retardation | Poor concentration and indecisiveness<br>Psychomotor retardation | difficult concentrating, difficult to concentrate, diminished ability to concentrate,<br>diminished ability to think, indecisiveness, poor concentration<br>psychomotor retardation |
| 3 | Loss of appetite and body weight | Loss of appetite and body weight | appetite decreased, decreased appetite, loss of appetite, weight loss |
| 4 | Increased appetite and body weight | Increased appetied and body weight | gain weight, increased appetite, weight gain |
| 5 | Insomnia | Insomnia | difficult falling asleep, disturbed sleep, early morning awakening, insomnia,<br>sleeplessness, wake up early |
| 6 | Loss of energy/fatigue | Loss of energy/fatigue | decreased activity, fatigue, feeling tired, feeling tired easily, loss of energy,<br>reduced energy, hypersomnia |
| 7 | Psychomotor agitation | Psychomotor agitation | agitated, agitation, irritability, psychomotor, agitation, restless |
| 8 | Suicidal tendency | Suicidal tendency | ideas of self harm, plans for suicide, SI, suicidal, suicidal ideation,<br>suicidal plan, suicide, suicide attempt, suicide thoughts |
| 9 | Psychotic symptoms | Delusion<br>Hallucination<br>Any mention of psychosis | Delude, delusion<br>psychosis<br>AH, hallucinate, hallucination, hallucinatory |
| 10 | Anxiety symptoms | Anxiety and stress | Anxious, apprehensive, distressed, nervous, stressful, tense, uptight, worried |
| 11 | Pain | Pain | Pain, H/A, headache |

## Appendix C Modeled propensity odds ratio and 95% confidence intervals, PCP subset

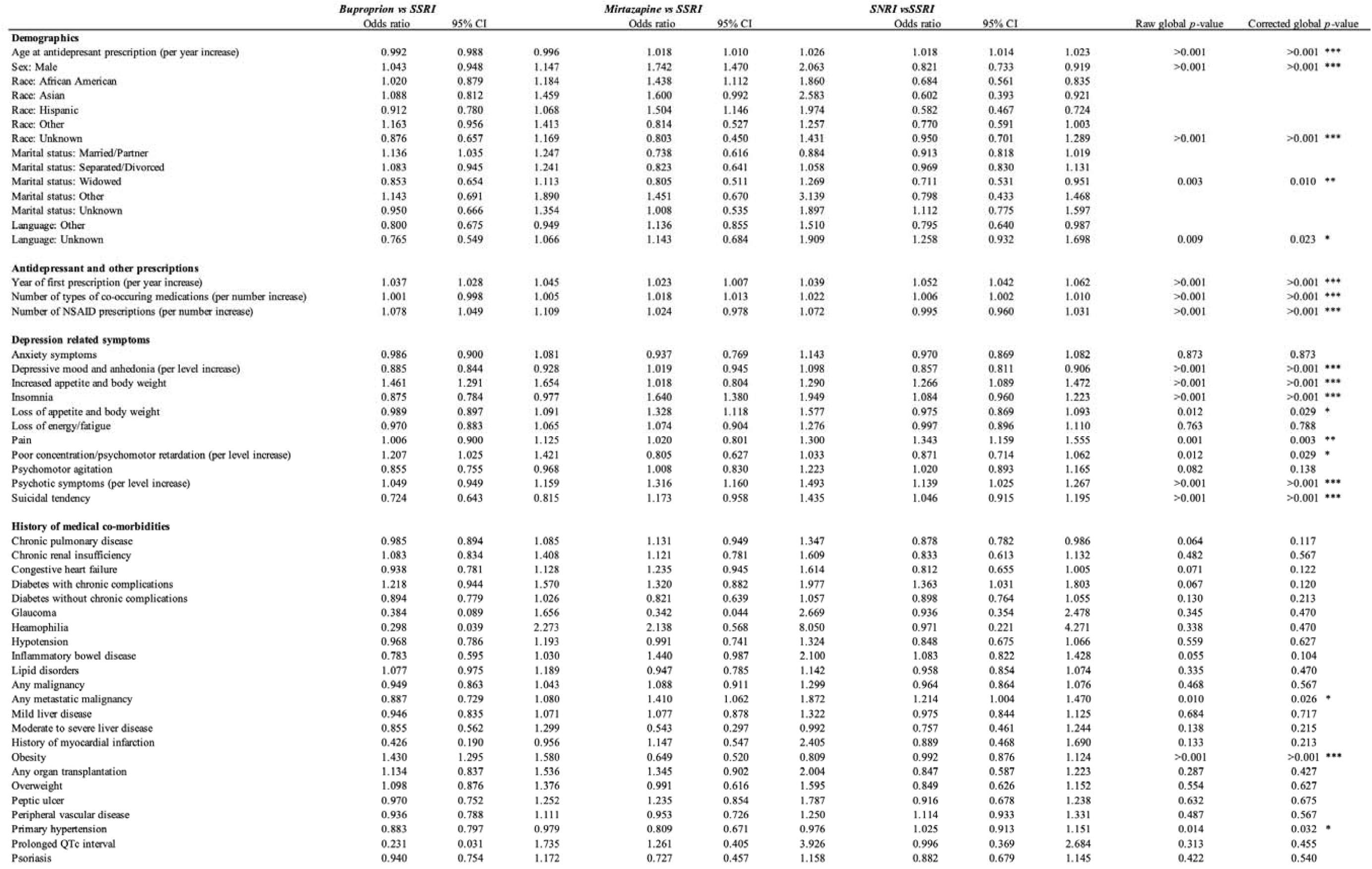
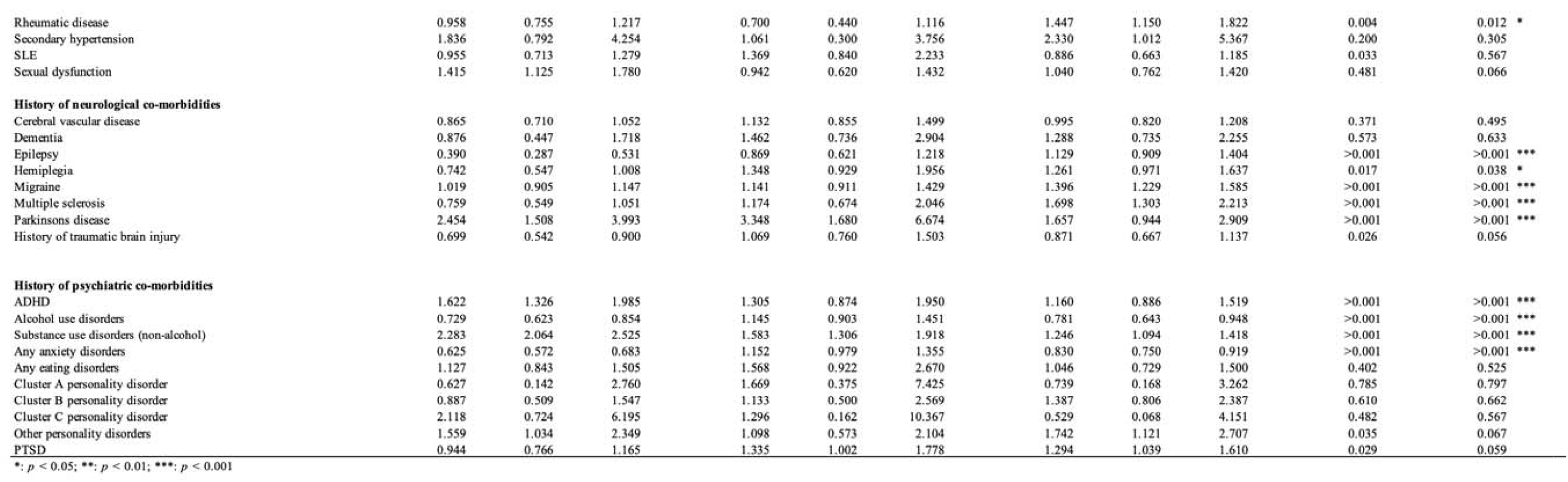

## Appendix D Forest plot for ORs and CIs of globally significant variables for multinominal regression model of antidepressant category propensity, PCP subset

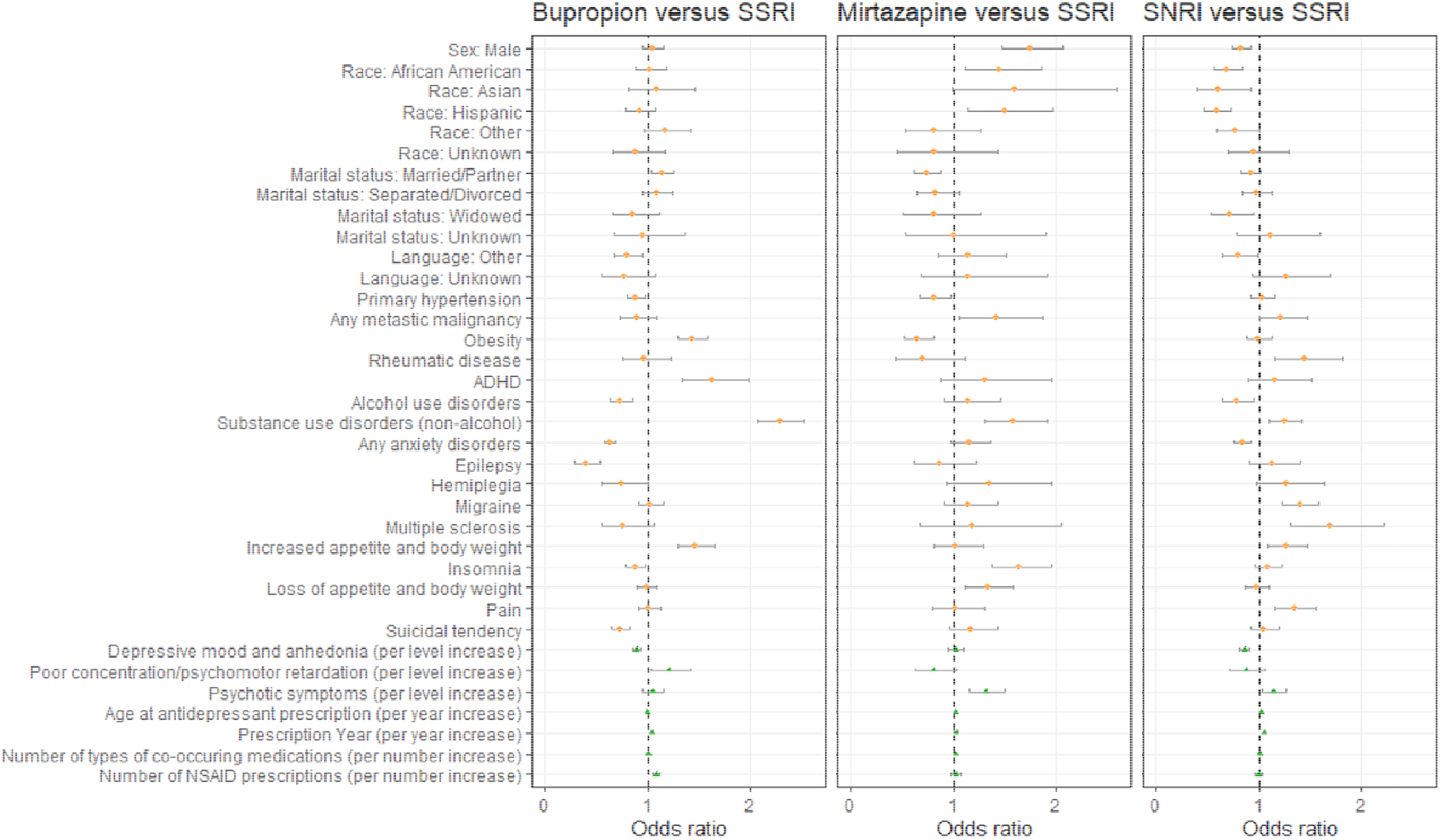

*Parkinson’s disease and Marital Status: Other are not shown for illustration purpose due to very wide confidence interval

## Appendix E Forest plot for ORs and CIs of of all variables for multinominal regression model of antidepressant category propensity, full data set

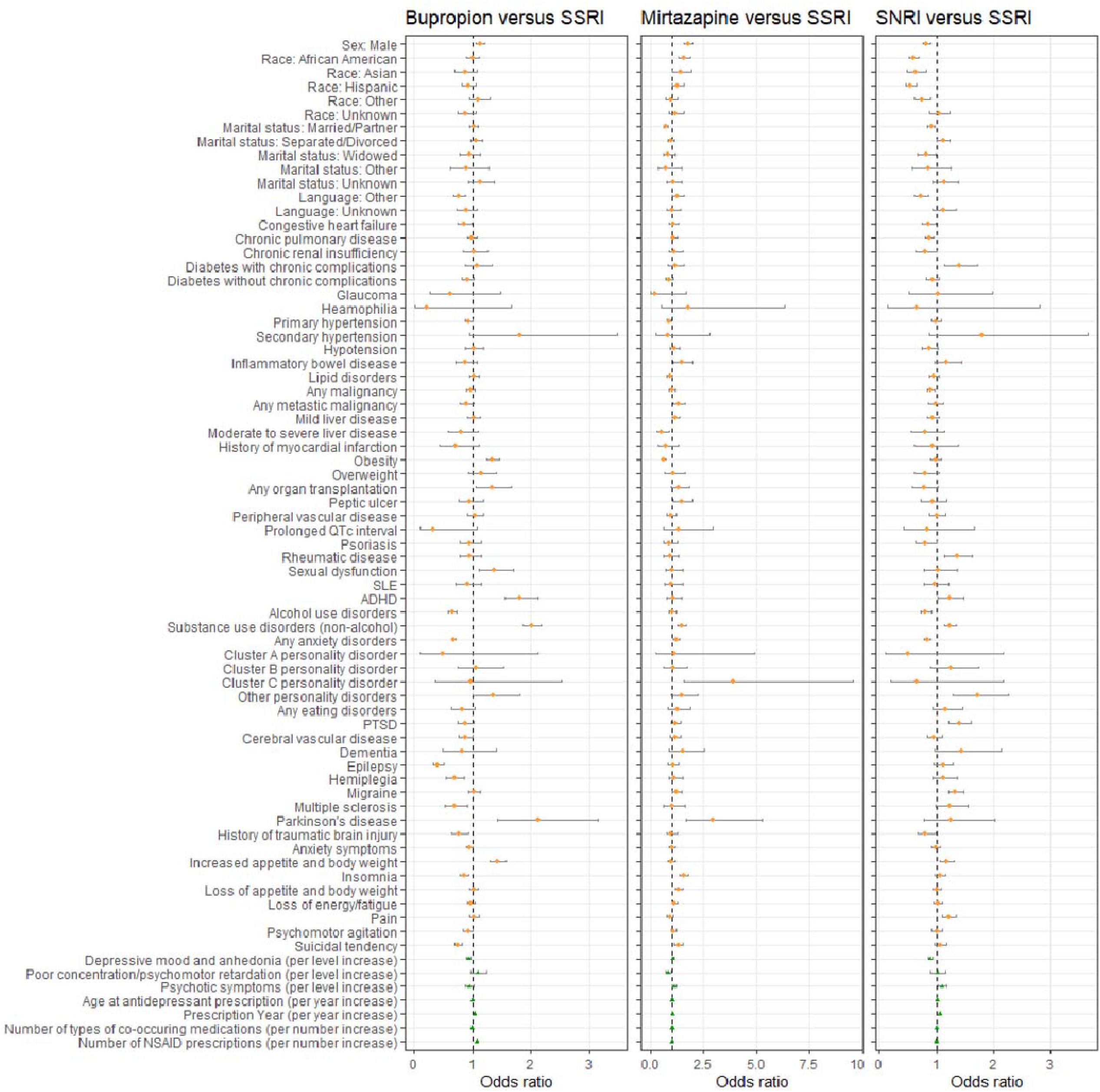

## Appendix F Forest plot for ORs and CIs of of all variables for multinominal regression model of antidepressant category propensity, PCP subset

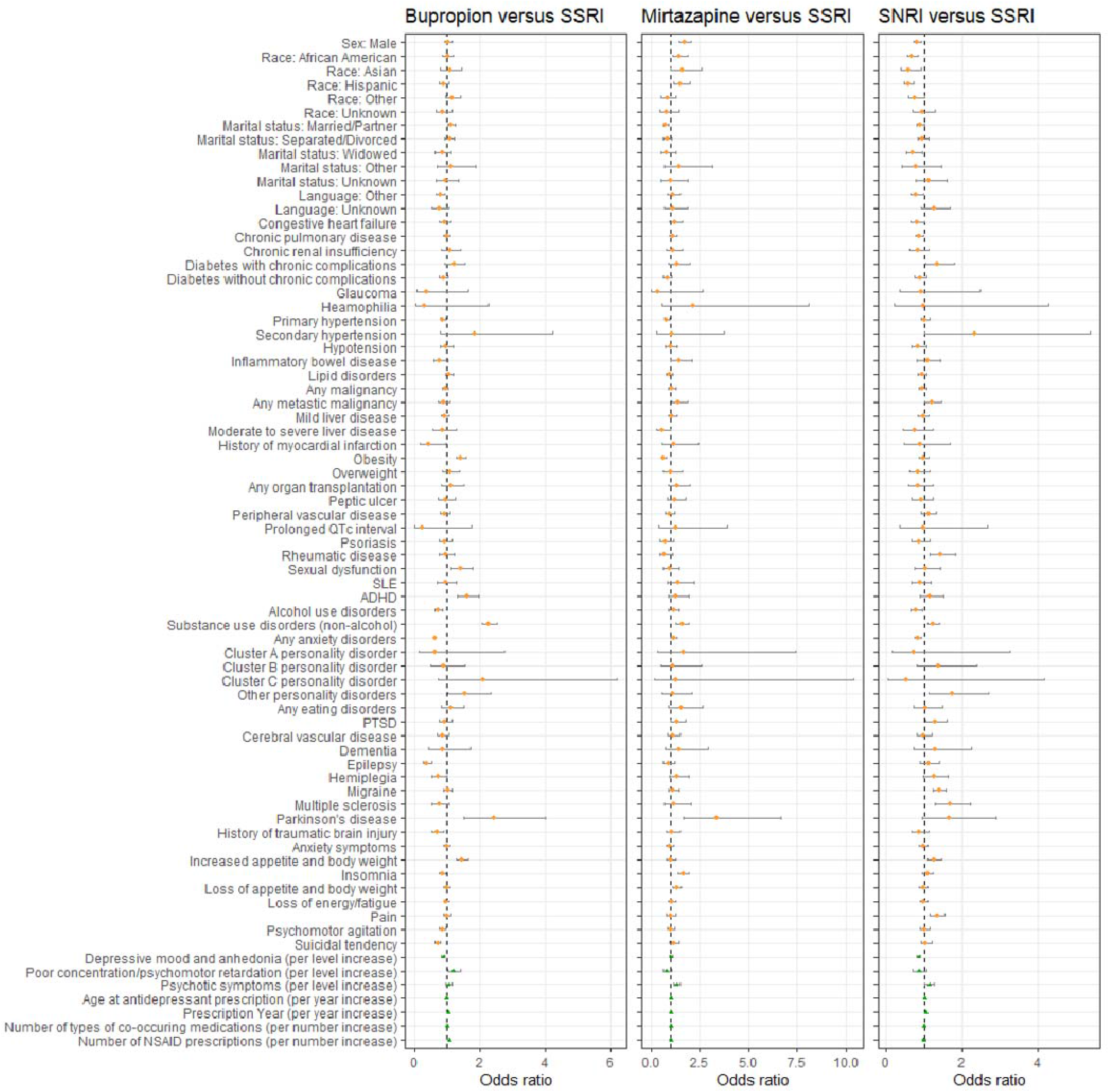

## Notes

### Funding Statement

No external funding was received for the purpose of this study.

### Author Declarations

This study is approved by the IRB of the MassGeneralBrigham (MGB) healthcare system (Boston, MA, USA; Protocol number: #2018P000765) which granted permission to the processing and analyses of the electronic health record data provided by MGB that were used for the purpose of this study.

## References

1. Hasin DS, Sarvet AL, Meyers JL, Saha TD, Ruan WJ, Stohl M, et al. Epidemiology of Adult DSM-5 Major Depressive Disorder and Its Specifiers in the United States. JAMA Psychiatry. 2018 Apr 1;75(4):336–46.

2. Bushnell GA, Stürmer T, Mack C, Pate V, Miller M. Who diagnosed and prescribed what? Using provider details to inform observational research. Pharmacoepidemiol Drug Saf. 2018 Dec;27(12):1422–6.

3. Gelenberg AJ, Markowitz JC, Rosenbaum JF, Thase ME, Trivedi MH, Rhoads RS. Practice Guideline for the Treatment of Patients with Major Depressive Disorder. American Psychiatric Association; 2010.

4. Sadock BJ, Sadock VA, Ruiz P. Kaplan & Sadock’s synopsis of psychiatryl_J: behavioral sciences/clinical psychiatry. Philadelphia: Lippincott Williams; 2015.

5. Stephen M. Stahl NM. Stahl’s Essential Psychopharmacology: Neuroscientific Basis and Practical Applications, 4th Edition. Cambridge University Press; 2013.

6. Hales RE, Yudofsky SC, Roberts LW. The American Psychiatric Association Publishing Textbook of Psychiatry. 6th ed. Washington DC: American Psychiatric Publishing; 2014.

7. MassGeneralBrigham Healthcare System. Research Patient Data Registry (RPDR) [Internet]. [cited 2018 Aug 1]. Available from: https://rc.partners.org/about/who-we-are-risc/research-patient-data-registry

8. Quan H, Sundararajan V, Halfon P, Fong A, Burnand B, Luthi J-C, et al. Coding algorithms for defining comorbidities in ICD-9-CM and ICD-10 administrative data. Med Care. 2005 Nov;43(11):1130–9.

9. Jette N, Beghi E, Hesdorffer D, Moshé SL, Zuberi SM, Medina MT, et al. ICD coding for epilepsy: past, present, and future--a report by the International League Against Epilepsy Task Force on ICD codes in epilepsy. Epilepsia. 2015 Mar;56(3):348–55.

10. National Lipid Association. Commonly Used Lipidcentric ICD-10 (ICD-9) Codes. 2015;

11. American Academy of Ophalmology. Glaucoma Quick Reference Guide. 2015;

12. The Web’s Free ICD-9-CM Medical Coding Reference [Internet]. 2019. Available from: http://www.icd9data.com/

13. The Web’s Free 2019 ICD-10-CM/PCS Medical Coding Reference [Internet]. 2019. Available from: https://www.icd10data.com/

14. Anglin R, Yuan Y, Moayyedi P, Tse F, Armstrong D, Leontiadis GI. Risk of upper gastrointestinal bleeding with selective serotonin reuptake inhibitors with or without concurrent nonsteroidal anti-inflammatory use: a systematic review and meta-analysis. Am J Gastroenterol. 2014 Jun;109(6):811–9.

15. Jiang H-Y, Chen H-Z, Hu X-J, Yu Z-H, Yang W, Deng M, et al. Use of selective serotonin reuptake inhibitors and risk of upper gastrointestinal bleeding: a systematic review and meta-analysis. Clin Gastroenterol Hepatol. 2015 Jan;13(1):42–50.e3.

16. Diagnostic and Statistical Manual of Mental Disorders, 5th Edition (DSM-5). American Psychiatric Association; 2013.

17. spaCy Website [Internet]. [cited 2019 Jan 8]. Available from: https://spacy.io/

18. Salk RH, Hyde JS, Abramson LY. Gender differences in depression in representative national samples: Meta-analyses of diagnoses and symptoms. Psychol Bull. 2017 Aug;143(8):783–822.

19. Wong J, Motulsky A, Abrahamowicz M, Eguale T, Buckeridge DL, Tamblyn R. Off-label indications for antidepressants in primary care: descriptive study of prescriptions from an indication based electronic prescribing system. BMJ. 2017 Feb 21;356:j603.

20. Waitzfelder B, Stewart C, Coleman KJ, Rossom R, Ahmedani BK, Beck A, et al. Treatment initiation for new episodes of depression in primary care settings. J Gen Intern Med. 2018 Aug;33(8):1283–91.

21. Johnson CF, Williams B, MacGillivray SA, Dougall NJ, Maxwell M. “Doing the right thing”: factors influencing GP prescribing of antidepressants and prescribed doses. BMC Fam Pract. 2017 Jun 17;18(1):72.

22. Zimmermann LL, Diaz-Arrastia R, Vespa PM. Seizures and the role of anticonvulsants after traumatic brain injury. Neurosurg Clin N Am. 2016 Oct;27(4):499–508.

23. Marrie RA, Reider N, Cohen J, Trojano M, Sorensen PS, Cutter G, et al. A systematic review of the incidence and prevalence of sleep disorders and seizure disorders in multiple sclerosis. Mult Scler. 2015 Mar;21(3):342–9.

24. Wang JZ, Vyas MV, Saposnik G, Burneo JG. Incidence and management of seizures after ischemic stroke: Systematic review and meta-analysis. Neurology. 2017 Sep 19;89(12):1220–8.

25. Perlis RH, Iosifescu DV, Castro VM, Murphy SN, Gainer VS, Minnier J, et al. Using electronic medical records to enable large-scale studies in psychiatry: treatment resistant depression as a model. Psychol Med. 2012 Jan;42(1):41–50.

26. Esteban S, Rodríguez Tablado M, Peper F, Mahumud YS, Ricci RI, Kopitowski K, et al. Development and Validation of Various Phenotyping Algorithms for Diabetes Mellitus Using Data from Electronic Health Records. Stud Health Technol Inform. 2017;245:366–9.

27. Beaulieu-Jones BK, Greene CS, Pooled Resource Open-Access ALS Clinical Trials Consortium. Semi-supervised learning of the electronic health record for phenotype stratification. J Biomed Inform. 2016 Dec;64:168–78.

28. van Eeghen C, Littenberg B, Holman MD, Kessler R. Integrating behavioral health in primary care using lean workflow analysis: A case study. J Am Board Fam Med. 2016 Jun;29(3):385–93.

